# Human metapneumovirus (hMPV) test-positivity sentinel laboratory trends in England, 2012 to 2025

**DOI:** 10.1101/2025.03.04.25322918

**Authors:** Mary A. Sinnathamby, Beth Savagar, Chris Rawlinson, Megan Rome, Alex Allen, Conall Watson

**Affiliations:** Respiratory Virus Section, Immunisation and Vaccine-Preventable Diseases Division, Public Health Programmes, UK Health Security Agency, London

**Author notes:** joint first and senior authors (Mary A. Sinnathamby, Beth Savagar, Alex Allen, Conall Watson).

## Abstract

Human metapneumovirus is a common upper respiratory tract infection, routinely monitored as part of laboratory respiratory virus surveillance in England. Given the recent international interest in trends of hMPV, we describe the current and historical trends of hMPV in England overall and by age group using activity threshold methods.

## Rationale

Human metapneumovirus (hMPV) is a common cause of upper respiratory tract infection and was first identified in 2001 by Dutch virologists studying unexplained respiratory infections in young children, which ranged from mild to severe cough, bronchiolitis and pnemonia (1–3). In England, hMPV activity has been routinely monitored as part of respiratory virus surveillance since the 2009 influenza A(H1N1)pdm09 pandemic through the Respiratory DataMart sentinel laboratory system (4). Seasonal epidemics of hMPV in England are typically observed in late winter and early spring (2,5).

Given recent interest in trends of hMPV cases, we describe the current and historical trends of hMPV in England using activity threshold methods to assess and hMPV levels (2,6–9).

## hMPV monitoring in England

hMPV activity is monitored through sentinel primary and secondary care laboratory surveillance systems in England (4,5). For this study, hMPV positivity (%) data was collated from the Respiratory DataMart secondary care sentinel laboratory system (4). Positivity was computed as the weekly number of laboratory-confirmed hMPV positive tests out of all samples tested for hMPV in a given week. We analysed data from week 27 2012 (when DataMart had reached system maturity) to week 2 2025, with each season represented as a 52 week period (e.g. week 27 year 1 to week 26 year 2), enabling assessment of seasonal trends in hMPV activity.

Overall (all-age) hMPV positivity (%) was computed, and by age group (0-4, 5-14, 15-44, 45-64 and 65+ years). To assess hMPV activity levels, moving epidemic method (MEM) thresholds were calculated for overall positivity (%) (7,9). For positivity by age group, mean standard deviation (MSD) thresholds were calculated as computing MEM thresholds by age groups resulted in low and moderate thresholds below the baseline threshold due to the wide variation in positivity by age group (8). Both the MEM and MSD thresholds were computed using five seasons of data from week 27 2016 to week 26 2024, excluding the COVID-19 pandemic seasons (week 27 2019 to week 26 2022) (7–9).

## Trends in hMPV

Between 2012/13 and 2023/24 (with the exception of the 2020/21 and 2021/22 seasons), overall hMPV positivity (%) has followed a consistent winter seasonality which coincides with the circulation of influenza and RSV each year. The 2020/21 saw a significant drop in hMPV positivity explained by the COVID-19 pandemic public health and social measures as observed with other seasonal respiratory viruses. The highest hMPV positivity (%) recorded in the data was observed during the 2021 to 2022 season, peaking at 10.33% in week 49, whichwas at a time when public health and social measures were eased during the COVID- 19 pandemic initiating a rise in cases again as observed with other respiratory viruses.The median peak positivity (%) for hMPV was 5.54% (interquartile range: 5.07-6.09%) (Figure 1). When assessing the timing of peaks between 2012/13 and 2023/24 (excluding the COVID- 19 pandemic seasons in 2020/21 and 2021/22), 60% (6 of 10 seasons) of peaks occurred in winter (ranging between week 52 to week 03) and 40% (4 of 10 seasons) of peaks occurred in spring (ranging between week 12 and 17) (Figure 1).

**Figure 1.**
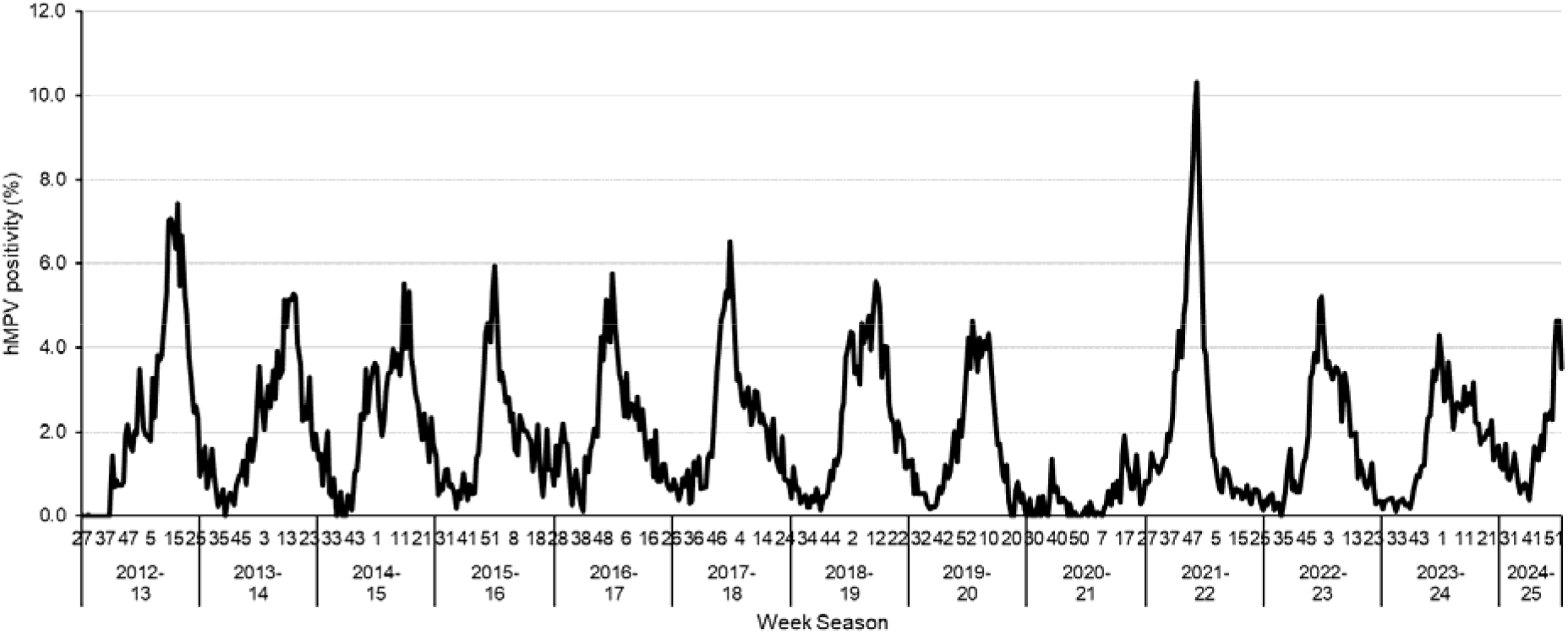
Overall hMPV positivity (%), week 27 2012 to week 2 2025

**Figure 1.**
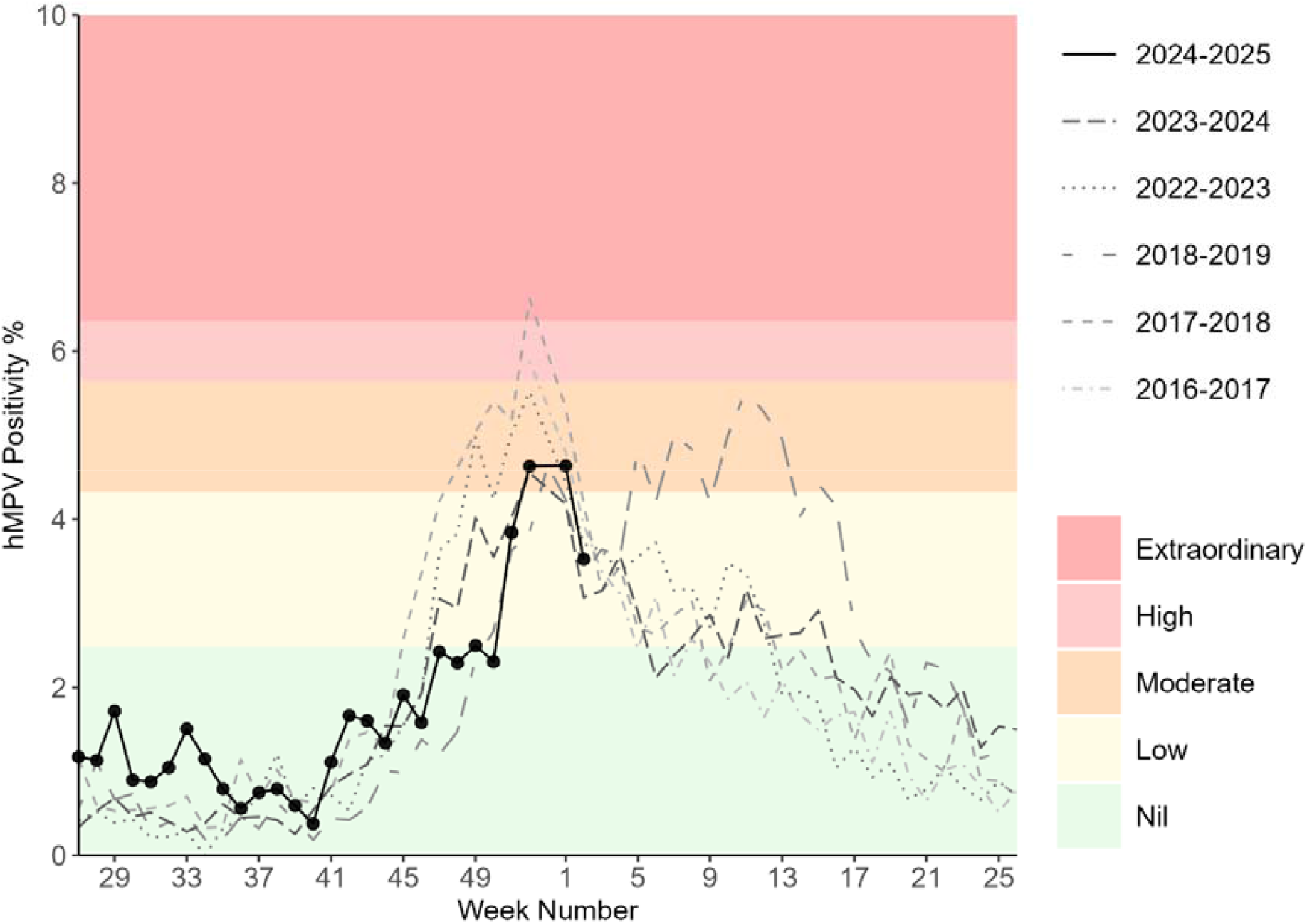
Overall (all ages combined) hMPV positivity (%) with MEM thresholds, week 26 2016 to week 2 2025

Overall hMPV positivity (%) during the 2024 to 2025 season has been consistent with historic trends with the highest positivity observed at 4/6% during week 52 2024 and week 1 2025 so far. This corresponded to moderate activity levels based on MEM calculations and is the same or lower than peak activity levels observed during previous seasons (Figure 2). A decline in week 2 2025 has been observed at 3.52%.

**Figure 2.**
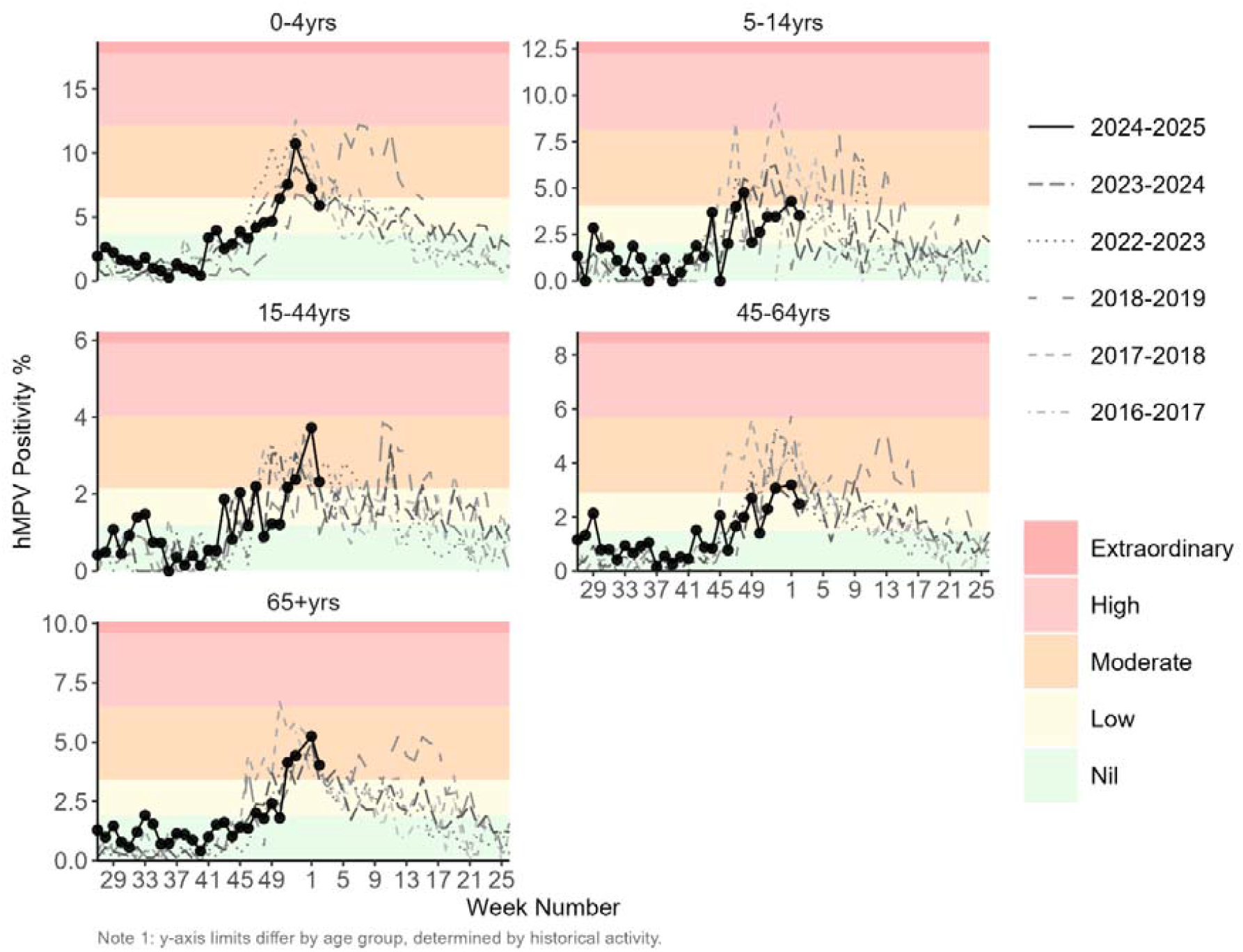
hMPV positivity (%) by age groups with MSD thresholds, week 26 2016 to week 2 2025

Similarly, hMPV positivity (%) by age group followed expected trends, with positivity (%) peaking at similar levels to previous seasons (Figure 3) and remaining within moderate activity levels across all age groups based on MSD calculations. As in previous seasons, the highest positivity (%) by age group was observed in 0-4 year olds, with a maximum positivity (%) of 10.73% in week 52 2024. This was consistent with the median peak positivity of 11.11% (IQR: 10.34 - 12.04, range: 8.89-12.59) observed in this age group between 2016/17 and 2023/24.

## Discussion

Our study of sentinel laboratory detection positivity provides evidence that the circulation of hMPV in England in 2024/25 has not differed from trends observed in previous historical seasons (2). To our knowledge, this is the first study to categorise hMPV activity using thresholds; which emphasises the moderate positivity levels of hMPV for this current season overall and across most age groups.

Furthermore, integrated routine surveillance for respiratory viruses is important to monitor that trends are circulating at expected levels in comparison to historical trends (10) to enable proportionate public health action and messaging. For example, the highest overall peak in hMPV positivity in England was noted in 2021/22 at a time when public health and social measures were lifted during the COVID-19 pandemic which inevitably saw a surge in positivity as observed with other respiratory viruses (11,12).

To conclude, our study highlights that hMPV activity in England remain at expected levels in the current 2024/25 season in comparison to previous seasons and has demonstrated the importance of differentiating activity levels to aid public health communication by using threshold settings methods, particularly when multiple respiratory viruses are circulating at the same time.

## Ethical statement

This work was conducted as part of routine surveillance and investigation activity, and thus ethical approval was not required.

## Use of artificial intelligence tools

None declared.

## Data availability

Data used in this study is available on publicly published national influenza and COVID-19 reports available from: <u>National flu and COVID-19 surveillance reports: 2023 to 2024 season - GOV.UK</u> (www.gov.uk)

Any other requests for data should be submitted to the UKHSA Office for Data Release, available from: <u>Accessing UKHSA protected data - GOV.UK</u> (www.gov.uk)

## Acknowledgments

We would like to thank all teams and organisations who contribute consistent data for the year around surveillance data for respiratory viruses.

## Authors’ contributions

MS and AA designed the study. MS and BS undertook the analyses. CR, MR and CW manage and provided data from the surveillance system used in this study.

